# Statins for Primary Prevention of Cardiovascular Disease in Germany: Benefits and Costs

**DOI:** 10.1101/2024.10.17.24315649

**Authors:** Alexander Dressel, Felix Fath, Bernhard K. Krämer, Gerald Klose, Winfried März

## Abstract

**Background:** The reduction of LDL cholesterol lowers the risk of coronary and cerebrovascular events in individuals without manifest cardiovascular diseases. In Germany, statins may only be prescribed at the expense of statutory health insurance for patients with atherosclerosis-related diseases or those at high cardiovascular risk (over 20 percent event probability within the next 10 years, calculated using one of the “available risk calculators”). However, international guidelines recommend lower risk thresholds for the use of statins.

**Methods:** The health and economic impacts of different risk thresholds for statin use in primary prevention within the German population are estimated for thresholds of 7.5, 10, and 15 percent over 10 years, based on the US *Pooled Cohort Equation* (PCE) which has been validated for Germany, using Markov models.

**Findings:** Cost-effectiveness increases with a rising risk threshold, while individual benefit decreases with age at the start of treatment. The use of statins at a risk of 7.5 percent or more is cost-effective at any age (cost per QALY between 410 and 2,100 euros). In none of the examined scenarios does the proportion of the population qualifying for statin therapy exceed 25 percent.

**Interpretation:** Lowering the threshold for statin therapy based on age to a risk of 7.5 percent, estimated with the US PCE, aligns statin prescription with international standards. There is no urgent rationale for applying age-stratified risk thresholds using the SCORE2 proposed for Europe, which tends to underestimate actual risks.

## Introduction

For around two decades, it has been established that lowering **low-density lipoproteins** (LDL) with statins can reduce the incidence of cardiovascular events even in clinically healthy, asymptomatic individuals.^1–10^

According to the Cholesterol Treatment Trialists’ (CTT) Collaboration, lowering the concentration of LDL cholesterol (LDL-C) by (absolute) 1.5 mmol/l (60 mg/dl) will reduce the rate of cardiovascular events by (relatively) around one third ^7^ . A recent meta-analysis showed a 25 percent reduction in the rate of serious cardiovascular events and an 11 percent reduction in overall mortality with statins as a substance class. Atorvastatin and rosuvastatin had greater effects. ^8^

In Germany, it was previously impossible to exploit the potential of statin therapy in cases of moderate cardiovascular risk. This is because currently, according to Annex III of the Medicines Directive (number 35) of the Joint Federal Committee, lipid-lowering drugs can only be prescribed at the expense of the statutory health insurance funds in cases of atherosclerosis-related vascular disease or in “primary prevention” in cases of high cardiovascular risk of over **20 percent event rate in 10 years**, calculated using “one of the available risk calculators. ^11^ We pointed out many years ago that the “available risk calculators” lead to different assessments of the risk. ^12^ This finding is consistent with other sources ^13^ and is partly due to the fact that the predicted clinical endpoints in the risk calculators can be narrowly or broadly defined.

Current guidelines consider the risk thresholds for statin use to be well below a 20 percent chance of fatal and non-fatal atherosclerosis-related events in 10 years. The *US Department of Veterans Affairs* and the *US Department of Defense* recommend that patients with a cardiovascular risk of more than 12 percent over 10 years, calculated using the *Pooled Cohort Equation* (PCE ^14^ ) or a low-density lipoprotein cholesterol >190 mg/dl or diabetes mellitus should be offered a moderate-dose statin. ^15, 16^ The *American College of Cardiology* and the *American Heart Association* suggest treatment with a moderate-dose statin from a risk of 7.5 percent. ^17, 18^ The *National Institute for Health and Care Excellence* (NICE) recommends 20 mg atorvastatin daily from a risk of 10 percent in 10 years, calculated using the QRISK3 algorithm. ^19^ The *European Society of Cardiology* specifies age-dependent risk thresholds. If there is neither diabetes mellitus, chronic kidney disease nor severe dyslipidemia, a “very high risk” is assumed for asymptomatic people aged 50 to 69 years if the SCORE2 (Systematic Coronary Risk Estimation 2) ^13^ is above 10 percent. ^20^ In the age group under 50 years, a “very high risk” is assumed from 7.5 percent, in the age group over 70 years only from 15 percent with reference to the high age and competing risks.

As a result of the very similar lowering of intervention thresholds worldwide, the Federal Joint Committee decided on June 25, 2024 to initiate an opinion procedure to amend Annex III of the Medicinal Products Directive, the subject of which is the review of the outdated risk threshold of 20 percent in 10 years for the prescription of lipid-lowering drugs (https://www.g-ba.de/downloads/40-268-10605/2024-06-25_AM-RL-III_SN_Nr-35_Lipidsenker_TrG.pdf).

In addition, there is a draft law that aims to enshrine the right to prescribe statins in accordance with the current EAS-ESC guidelines if age-dependent risk thresholds of 7.5, 10 and 15 percent, calculated using the SCORE2 calculator, are exceeded (https://www.bundesgesundheitsministerium.de/fileadmin/Dateien/3_Downloads/Gesetze_und_Verordnungen/GuV/G/GHG_RefE_bf.pdf).

The aim of this study is to model the effects of different risk thresholds for the use of statins on the long-term prognosis of the German population, taking health economic aspects into account.

## Method

We developed Markov models simulating rates of cardiovascular events of 7.5, 10, and 15 percent over 10 years and associated costs and benefits for individuals with no prior history in relation to age at treatment initiation (25, 40, 50, 60, and 70 years) up to age 99 years. The health states included in the models were the occurrence of non-fatal and fatal coronary heart disease, the occurrence of non-fatal and fatal cerebrovascular events as assessed by the PCE ^14^ and death from other causes; for transitions and their probabilities, e.g. Table 1.

**Table 1.**
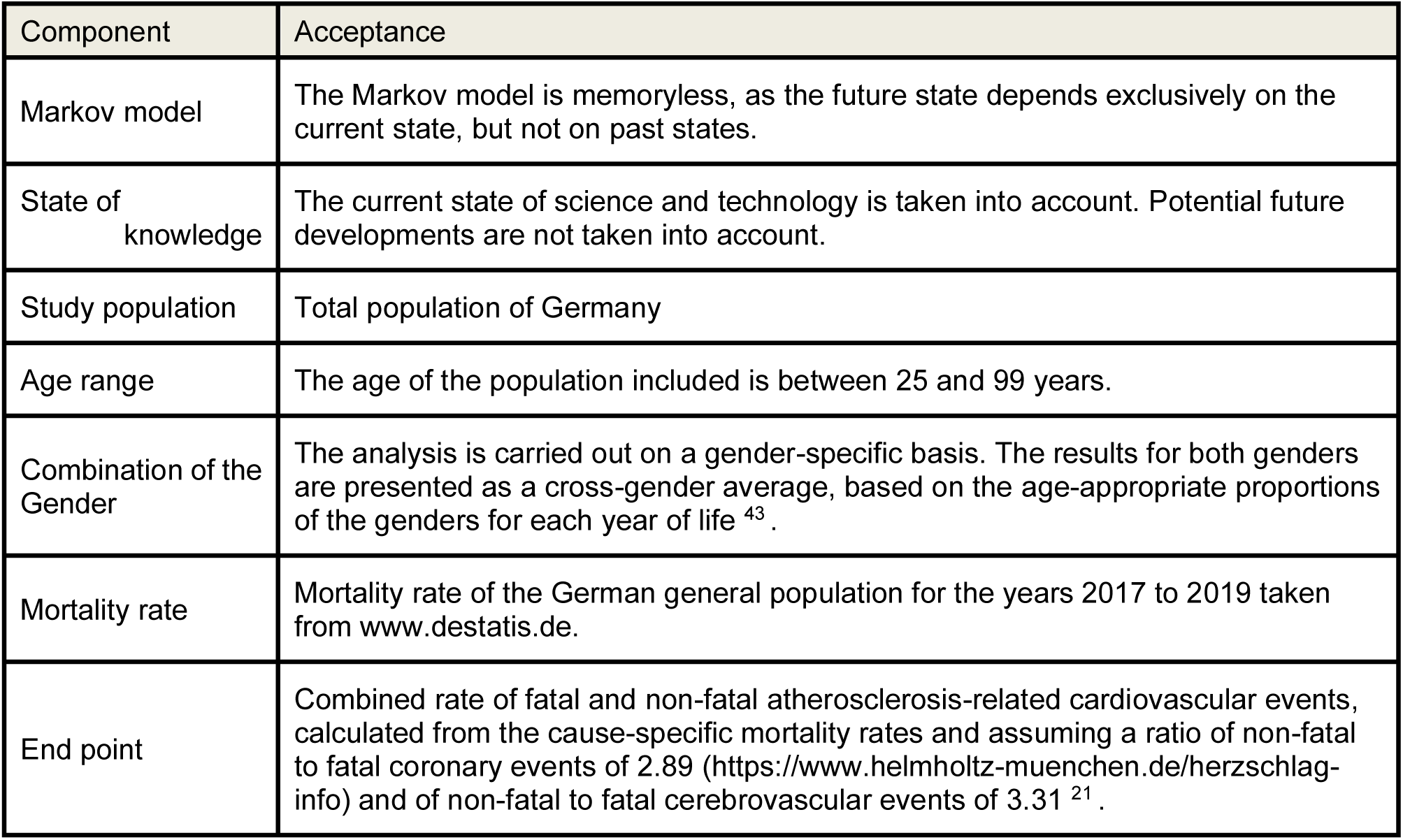
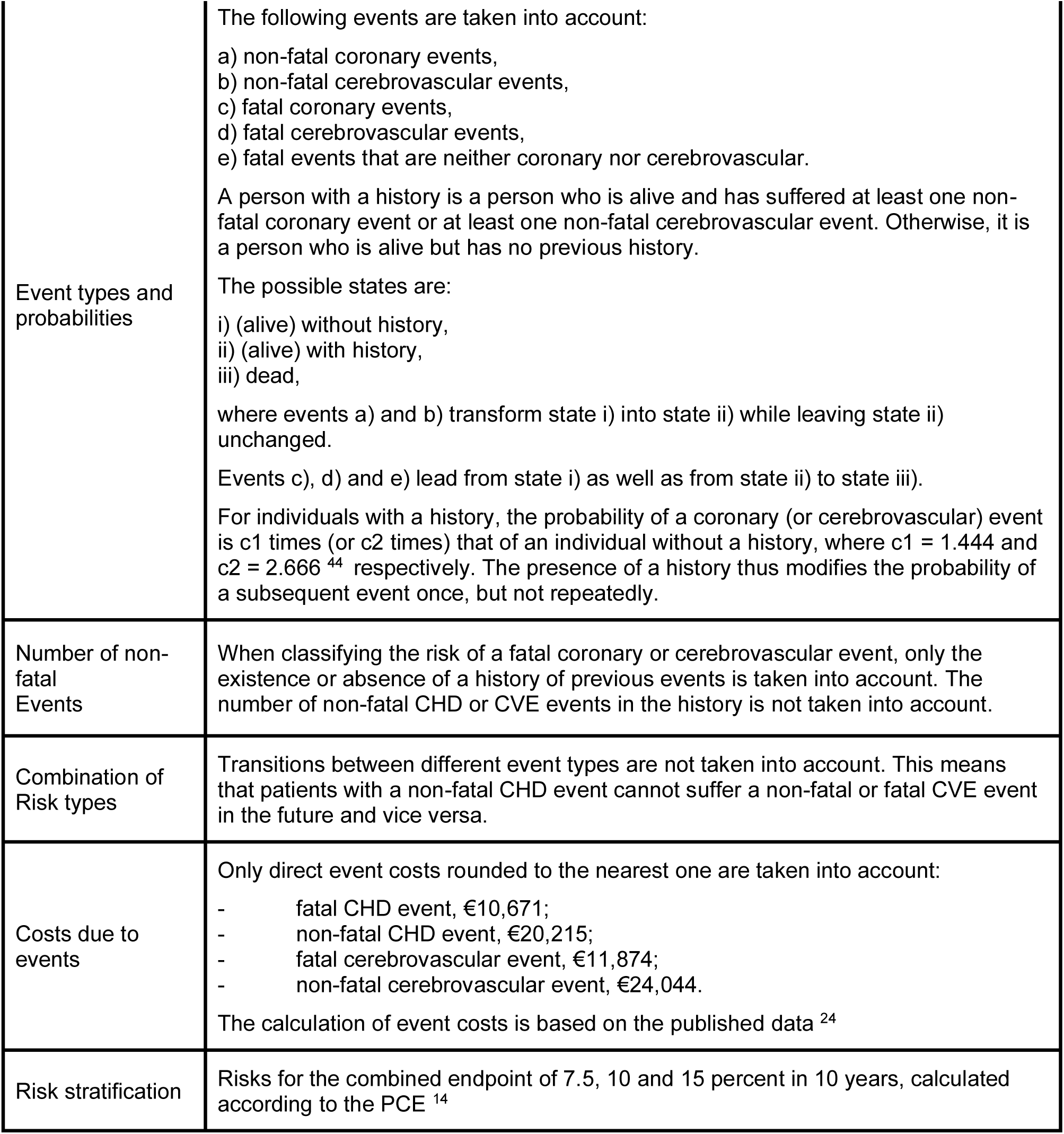
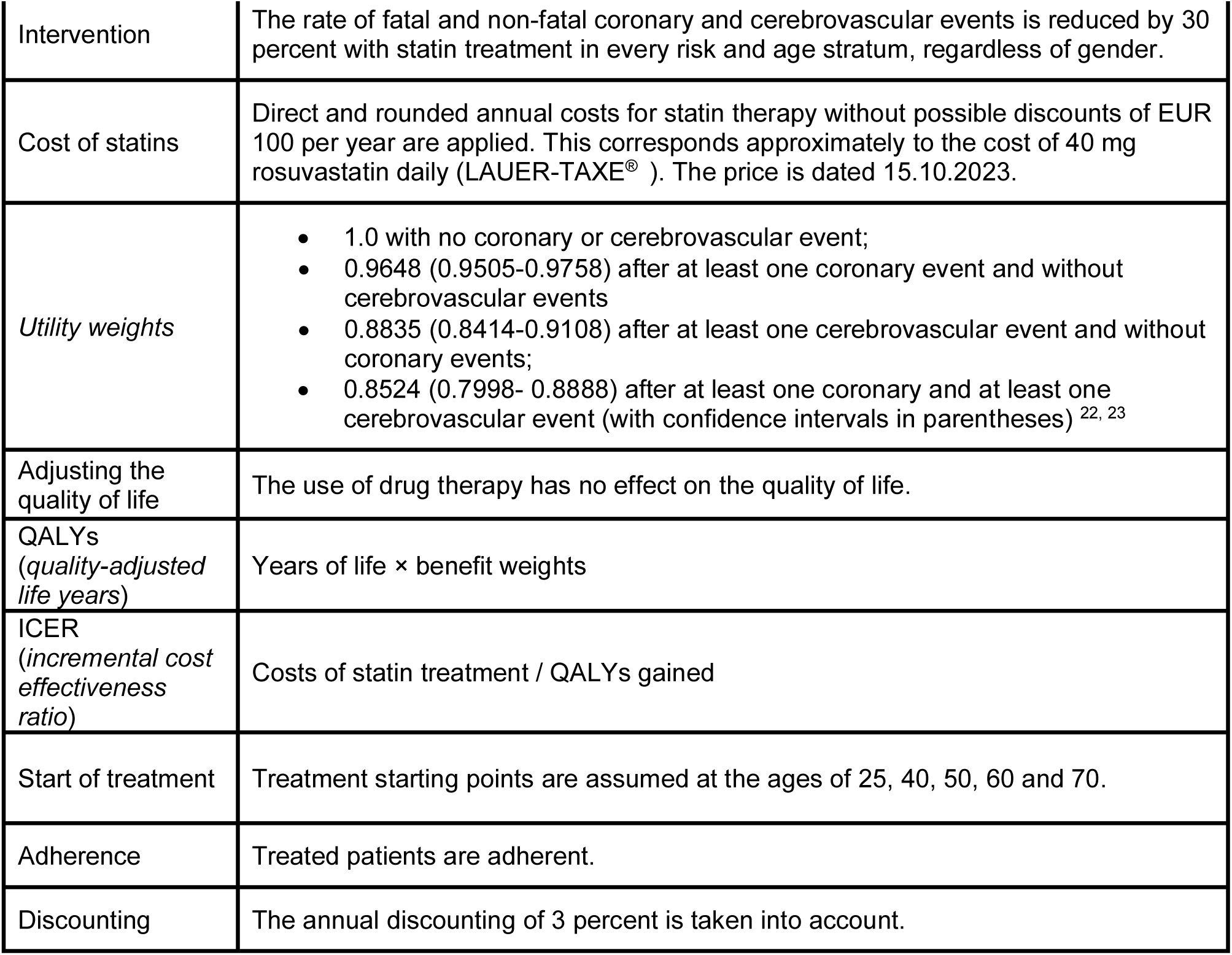
Model components and assumptions.

The calculation of the probabilities of fatal events is based on the mortality rates in the German general population stratified by age cohort and cause of death (www.destatis.com). In order to exclude special effects due to the coronavirus pandemic, we selected data from the period 2017 to 2019. To calculate the risks of non-fatal coronary and cerebrovascular events, we used a ratio of non-fatal to fatal events of 2.89 (https://www.helmholtz-muenchen.de/herzschlag-info) and 3.31. ^21^ Event rates were simulated using Markov chain Monte Carlo (MCMC) methods with a cycle length of one year and 20,000 bootstrap iterations up to a maximum age of 99 years (Figure 1).

**Figure 1.**
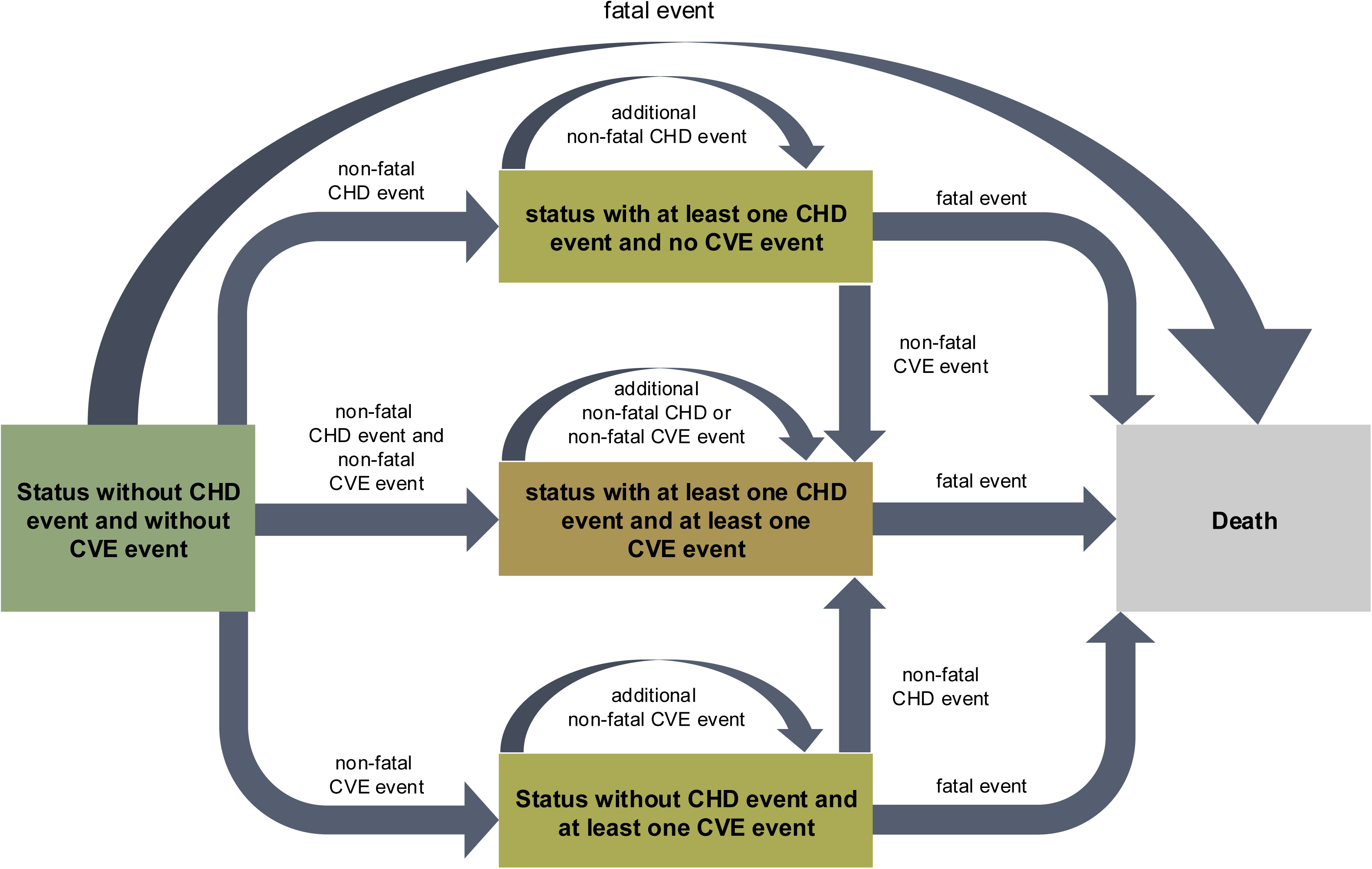
Cohort state transition model for clinically asymptomatic individuals. The occurrence of events was calculated using the Markov chain Monte Carlo method (MCMC) with a cycle length of one year and 20,000 *bootstrap* iterations up to a maximum age of 99 years. The transition probabilities were estimated from the mortality rates stratified by age cohort and cause of death using fixed ratios of non-fatal to fatal coronary and cerebrovascular events and adjusted to the survival data of the German general population available at www.destatis.com. In addition, the theoretical effects of lipid-lowering treatment with statins were taken into account. Non-fatal coronary and cerebrovascular events may coincide within any one-year period. Only one non-fatal coronary or cerebrovascular event is included in the calculation of the additional risk of patients with a history of coronary or cerebrovascular events; whether such an event occurs once or more frequently has no influence on the additional risk.

For different scenarios, we estimated the number of people needed to treat to prevent an event (numbers needed to treat, NNTs), based on a treatment period of 5 years, the quality-adjusted life years (QALYs) gained per person, savings from the avoidance of coronary and cerebrovascular events, incremental cost-effectiveness ratios (ICERs), the total costs of implementing these scenarios (budget impact) and their effects on mortality in the German population. We assumed a relative risk reduction of 30 percent through treatment with statins.

The *utility weights* were derived from the literature (Table 1). ^22, 23^ The estimation of direct medical costs is based on published data. ^24^ The cost of statin therapy was conservatively assumed to be 100 euros per year. Both costs and benefits were discounted at an annual rate of 3 percent to discount future values to today’s value. Indirect costs and benefits were not taken into account (Table 1).

All analyses were conducted separately for men and women; the costs and the QALYs and life years gained were calculated as cross-gender averages in order to derive the ICERs in particular. The proportions of men and women in the German population in 2022, stratified by age cohort, were used as a basis (www.destatis.com).

The Markov model was implemented using the software R (version 4.2.2).

## Results

The survival curves in Figure 2 A to D show the percentage of people among the living who suffer at least one coronary or cerebrovascular event over time for the scenarios of starting therapy at the age of 40, 50, 60 and 70 years and the 10-year risks of 7.5, 10 and 15 percent if there was no history of coronary or cerebrovascular events before starting therapy. As expected, the proportion of patients increases with age. The figure also illustrates the effects of lipid-lowering intervention with statins on the risks in the age strata.

**Figure 2.**
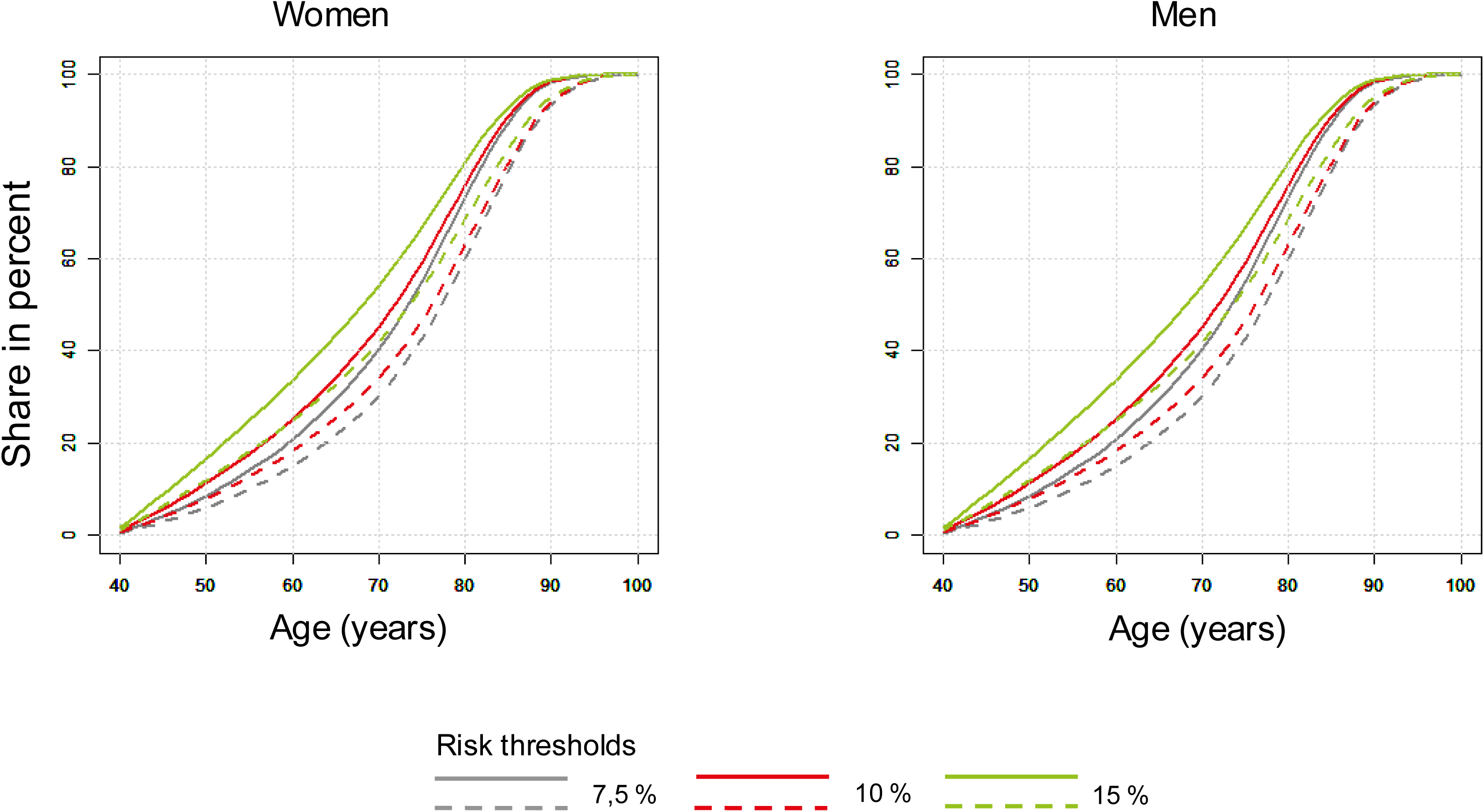
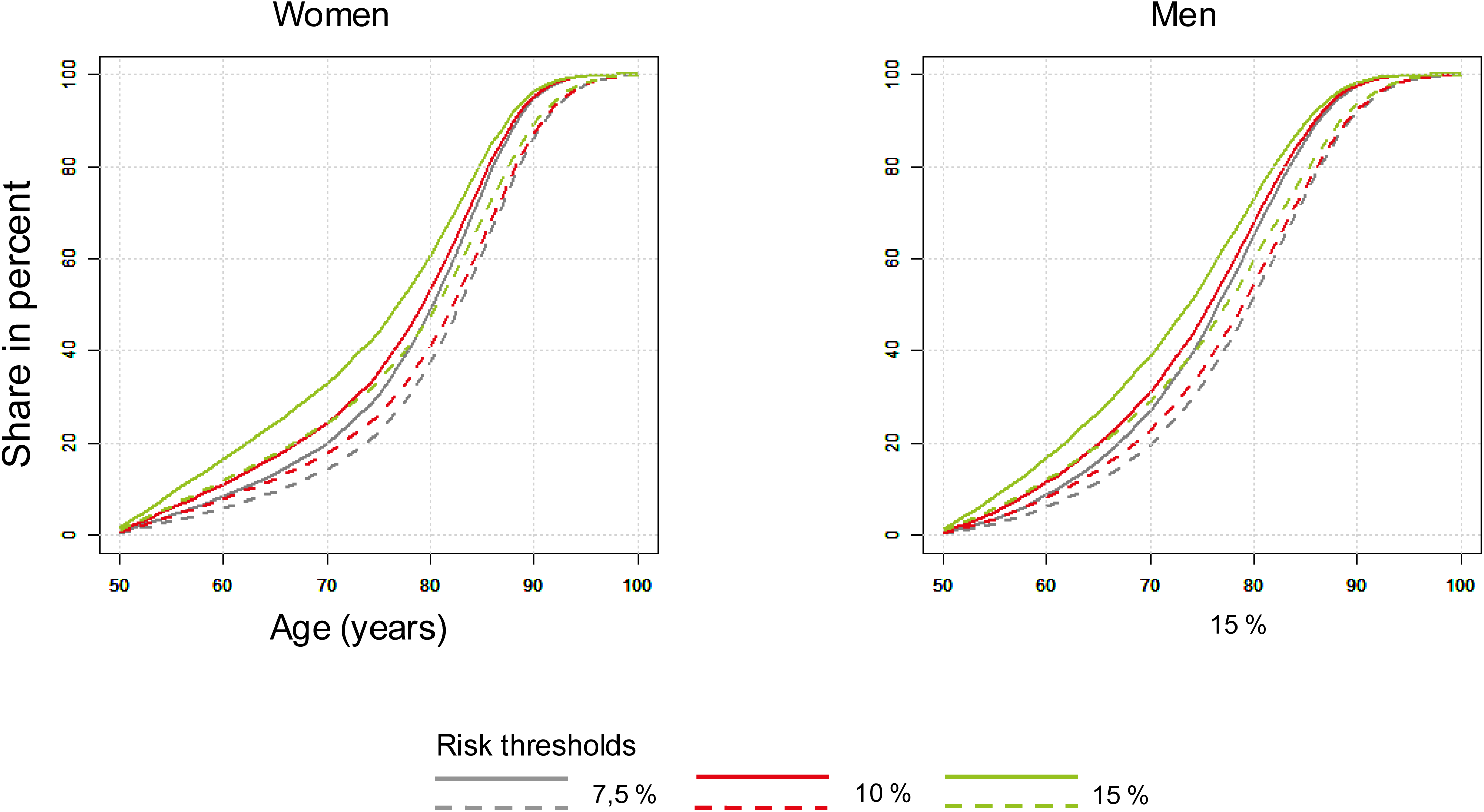
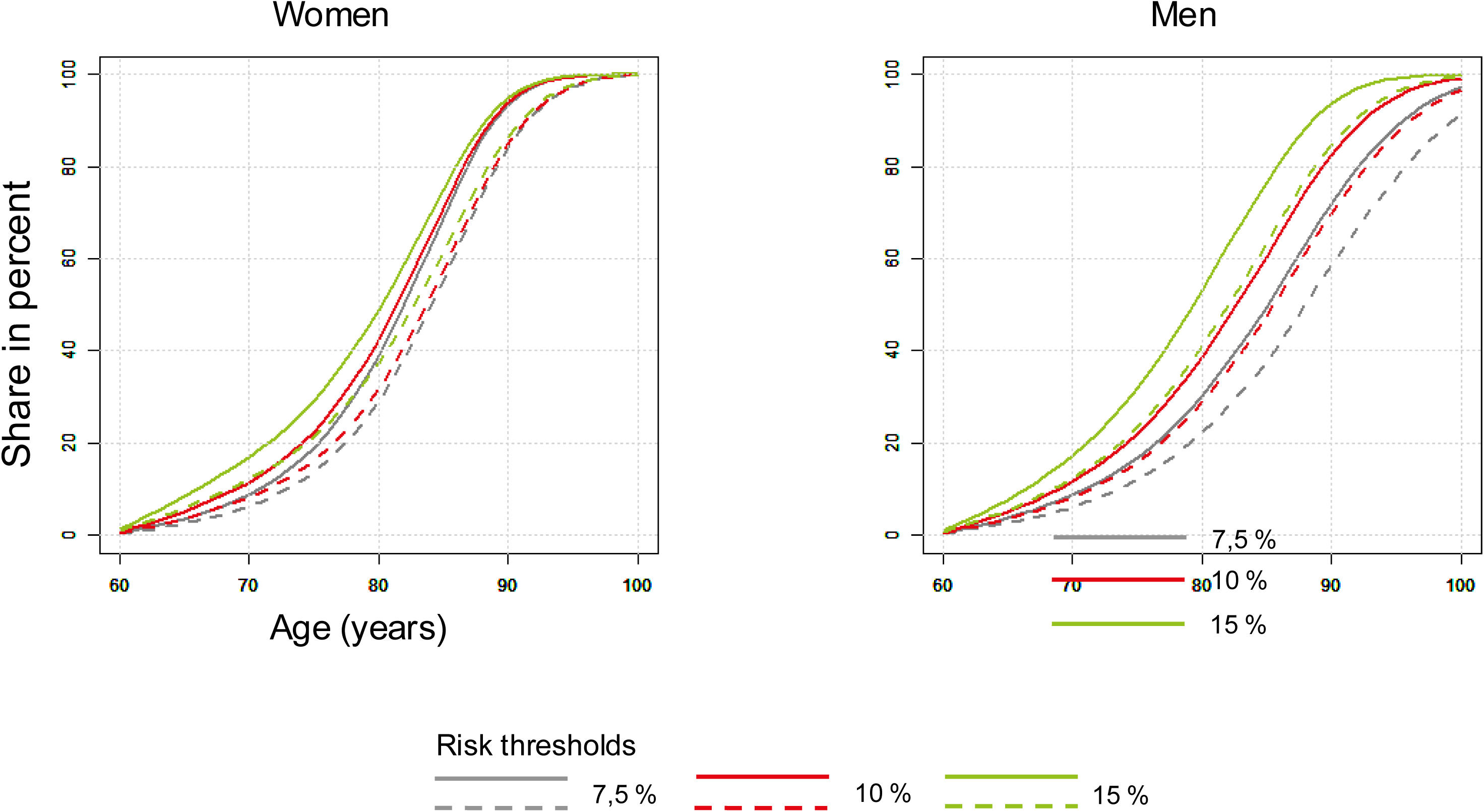
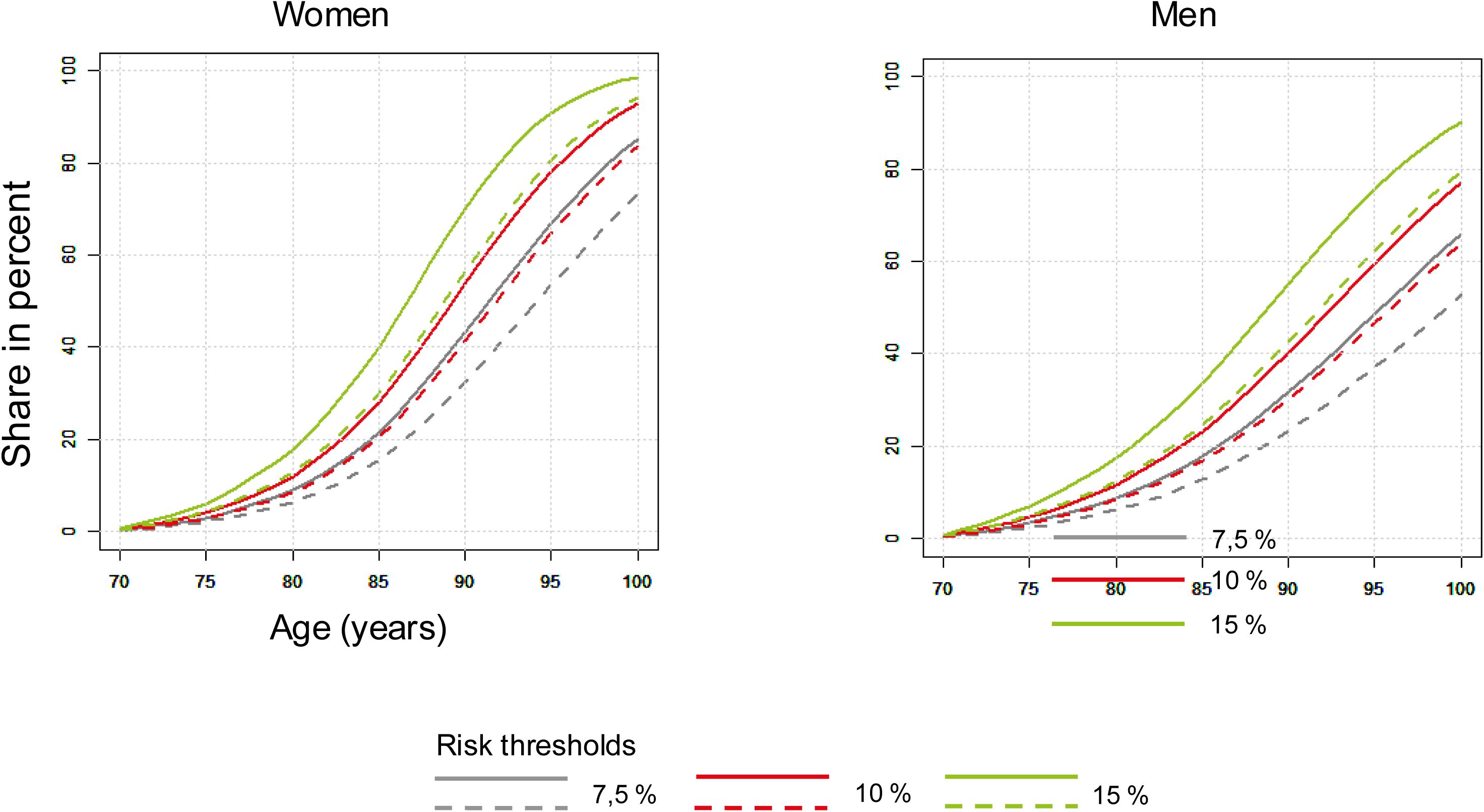
Estimated percentage of persons among the living with coronary or cerebrovascular events over time. Inclusion condition: no history of coronary or cerebrovascular events. The *solid* lines show the percentage of untreated living individuals with at least one coronary or cerebrovascular event in the population strata, the *dashed* lines show the percentage of statin-treated and living individuals. 10-year risks at the start of treatment 7.5 (*blue*), 10 (*red*) and 15 (*green*) percent. Start of treatment at the earliest age of 40 (*2A*), 50 (*2B*), 60 (*2C*) or 70 (*2D*) years. *Left:* women; *right:* men.

We have based the *numbers needed to treat* (NNTs) on 5 years, as this corresponds approximately to the duration of most prospective, randomized and placebo-controlled studies with statins. Table 2 shows that the NNTs decrease parallel to the risk threshold. In contrast, they increase when treatment is started later in life. The lowest NNT (24 in 5 years) is achieved with a risk threshold of 15 percent and treatment starting at the age of 25, the highest with a risk threshold of 7.5 percent and treatment starting at the age of 70 (NNT 429).

**Table 2.** Effects and costs of reducing the risk of fatal and non-fatal coronary and cerebrovascular events by 30 percent in Germany.

|  | Age at start of therapy (years) | NNT* (5 years) | QALYs gained with 95% CI (years) | ICERs (€ 1,000) | Avoided event costs discounted (€ 1,000) | Share of total population (%) | Mean risk of the treated population | Annual budget (€ billion) | Increase in life expectancy, treated population (years) | Increase in life expectancy, total population (years) |
| --- | --- | --- | --- | --- | --- | --- | --- | --- | --- | --- |
| 7.5 | 25 | 48 | 5.19 (4.80; 5.57) | 0.41 | 1.51 | 23.20 | 11.72 | 1.93 | -* | -* |
| 7.5 | 40 | 58 | 3.07 (2.78; 3.36) | 0.64 | 1.87 | 22.98 | 17.57 | 1.91 | 1.29 | 0.30 |
| 7.5 | 50 | 194 | 1.43 (1.21; 1.65) | 1.31 | 2.14 | 21.97 | 16.22 | 1.83 | 1.17 | 0.26 |
| 7.5 | 60 | 372 | 0.59 (0.39; 0.78) | 2.78 | 2.50 | 16.36 | 18.00 | 1.36 | 0.79 | 0.13 |
| 7.5 | 70 | 429 | 0.36 (0.20; 0.52) | 3.62 | 1.38 | 7.10 | 29.30 | 0.59 | 0.34 | 0.02 |
| 10 | 25 | 36 | 5.68 (5.30; 6.06) | 0.35 | 1.55 | 19.16 | 13.53 | 1.59 | -* | -* |
| 10 | 40 | 41 | 3.68 (3.39; 3.98) | 0.51 | 1.96 | 19.02 | 22.56 | 1.58 | 1.41 | 0.27 |
| 10 | 50 | 77 | 2.06 (1.83; 2.29) | 0.87 | 2.12 | 18.41 | 21.82 | 1.53 | 1.32 | 0.24 |
| 10 | 60 | 276 | 0.77 (0.58; 0.95) | 2.10 | 3.00 | 14.59 | 21.23 | 1.21 | 0.96 | 0.14 |
| 10 | 70 | 318 | 0.42 (0.26; 0.58) | 3.05 | 1.65 | 6.71 | 30.60 | 0.56 | 0.43 | 0.03 |
| 15 | 25 | 24 | 6.22 (5.86; 6.58) | 0.28 | 1.59 | 13.85 | 16.60 | 1.15 | -* | -* |
| 15 | 40 | 26 | 4.33 (4.04; 4.62) | 0.38 | 2.13 | 13.77 | 29.31 | 1.15 | 1.56 | 0.21 |
| 15 | 50 | 36 | 2.82 (2.58; 3.06) | 0.58 | 2.07 | 13.50 | 30.08 | 1.12 | 1.51 | 0.20 |
| 15 | 60 | 219 | 0.96 (0.78; 1.14) | 1.63 | 2.79 | 11.50 | 28.68 | 0.96 | 1.25 | 0.14 |
| 15 | 70 | 208 | 0.61 (0.45; 0.76) | 2.11 | 2.25 | 5.94 | 34.20 | 0.49 | 0.62 | 0.04 |

\* The data basis is not sufficient to calculate these values

For people with a risk of 7.5 percent in 10 years at the age of 25, a lifelong risk reduction of 30 percent (with a statin) leads to a gain of 5.19 QALYs; 410 euros must be spent for one QALY. In this scenario, around 23 percent of the population aged 25 years or older have a risk of more than 7.5 percent; 77 percent of the population are not treated (Table 2). The proportion of the total population receiving treatment only decreases significantly if the start of treatment is shifted to the sixtieth year of life or older, as only a few young people exceed the risk threshold of 7.5 percent. The proportion of people receiving treatment only falls below 10 percent of the population when treatment is started from the age of 70 (Table 2).

The cost-effectiveness increases as expected if the risk threshold is raised to 10 percent (in 10 years). A 25-year-old person then gains slightly more QALYs (5.68) compared to the same person with a risk of 7.5 percent (5.19 QALYs). The ICER is reduced from 410 to 350 euros. The proportion of people in need of treatment (10 percent or more risk, 25 years and older) in the total population is 19 percent. This only decreases significantly from the age of 60 onwards.

There is an even greater individual benefit for people with a risk of 15 percent and above: If risk reduction is started at the age of 25, a gain of 6.22 QALYs can be expected at a cost per QALY of only 280 euros. Around 14 percent of the population will be treated and therefore 86 percent will be excluded from treatment.

Remarkably, the absolute benefit of the risk reduction diminishes in parallel with the age at the start of treatment. At the age of 70, with a risk threshold of 7.5 percent, a gain of only 0.36 QALYs can be expected at a cost of 3620 euros per QALY. With risk thresholds of 10 and 15 percent, the gains are 0.42 and 0.61 QALYs, and incremental costs of EUR 3050 and EUR 2110 are incurred. The diminishing benefit of treatment arises from *competing risks*, the magnitude of which we have shown as the ratio of cardiovascular deaths to deaths from non-cardiovascular causes in Figure 3. For women aged 30, this ratio is around 50 and falls to 0.5 by the age of 80. For men aged 30, it is around 30 and also decreases to around 0.5 with age.

**Figure 3.**
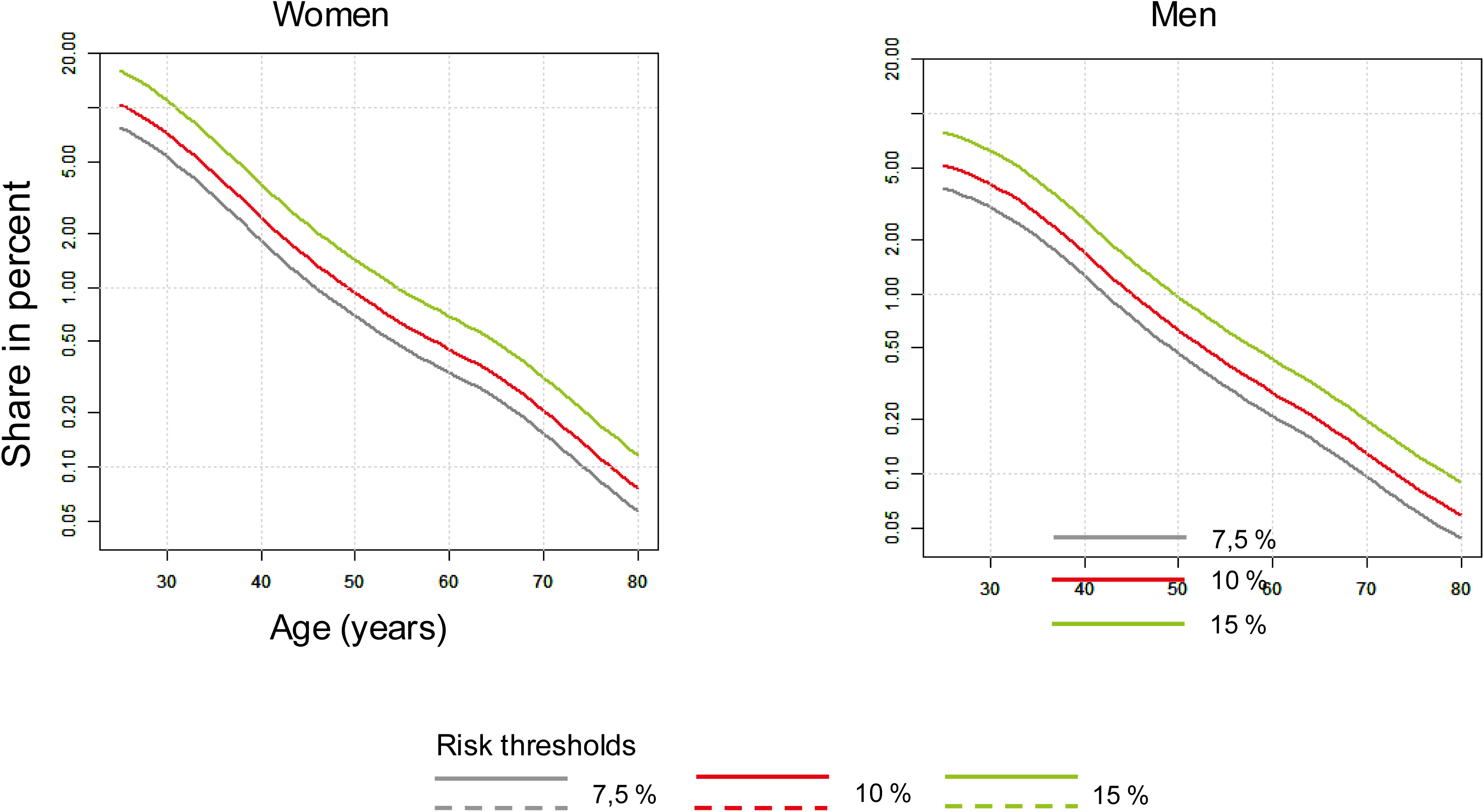
Influence of age on the ratios of the 10-year risk of fatal coronary or cerebrovascular events to the 10-year risk of death from other causes. The ordinate is logarithmically scaled. The lines show the progression of the ratios in the population strata with 10-year risks from 7.5 (*blue*), 10 (*red*) and 15 (*green*) percent for cardiovascular events calculated according to the *Pooled Cohort Equation* (PCE). *Left:* women; *right:* men.

### Effects on the budget (*budget impact*)

The costs from the perspective of the payers depend on the choice of risk threshold and the time of treatment start. They range between EUR 0.49 billion (risk threshold of 15 percent and start of treatment from the age of 70) and EUR 1.93 billion (risk threshold of 7.5 percent and start of treatment from the age of 25). For all risk thresholds, treatment costs only fall significantly when treatment begins at the age of 60 or later.

The treatment costs are offset by an increase in average life expectancy both in the treated group of people and in the population as a whole. Assuming treatment begins at the age of 40 and a risk threshold of 7.5%, this amounts to 1.29 years in the treated population (23% of the total population) and 0.30 years in relation to the total population. The increase in the average life expectancy of a treated population consisting of people over a certain age is lower than the gain in QALYs of an individual who starts treatment at a certain age. This is because starting treatment at a certain age implies that all older people with higher risks are also treated. This is illustrated by looking at the mean risk of the population to be treated (Table 2, column 9).

## Discussion

We have modeled the potential effects of primary prevention with statins based on the available evidence from randomized, placebo-controlled intervention trials and against the background of the current German healthcare landscape. In Germany, costs per QALY of less than €40,000 (roughly equivalent to gross domestic product per capita) are considered “very cost-effective” ^25^ costs per QALY of up to 120,000 euros are still considered cost-effective. In all scenarios examined, the prescription of statins in primary prevention would be highly cost-effective from a risk threshold up to the “most unfavorable” constellation, namely statin therapy from a risk threshold of 7.5 percent in 10 years from the age of 70. There is therefore no compelling justification, at least for statin therapy, to make the risk threshold dependent on age, as proposed by the ESC without explicit justification. ^20^ Based on our results, the competing risks increase almost 100-fold compared to an age of 30 years. The concept of competing risks implies that the risk of a particular event, in this case death from cardiovascular causes, is reduced by the increase in other competing causes of death. These considerations provide an unambiguous rationale for the ESC’s recommendation to raise the risk threshold for initiating statin therapy in parallel with age. It would possibly also be justified for significantly more costly treatment approaches than statin therapy. However, since starting statin therapy in older people at the risk threshold of 7.5 percent can still be classified as “very cost-effective”, we advocate leaving it at the low risk threshold of 7.5 percent in this group as well. This is because stratification by age would probably create unnecessary confusion rather than clarity in practice.

Our findings are based on the assumption that the cardiovascular risk is reduced by 30 percent through statin therapy. This requires a long-term reduction in LDL-C of 1.5 mmol/l (60 mg/dl). With an average LDL-C of around 3.34 mmol/l (129 mg/dl) in the German population ^26^ this corresponds to a relative reduction of 45 percent. This can be achieved with intensive statin monotherapy ^27^ but requires long-term adherence. However, our sensitivity analysis shows that even less optimistic assumptions hardly worsen the cost-effectiveness of statin therapy.

The cost-effectiveness of low-threshold and early statin therapy is also significantly better than that of other widely accepted non-medical life-saving measures. For example, the average cost of a QALY saved by preventing workplace exposure to pollutants is reported to be around USD 1.4 million, while the cost of containing pollutants in the environment is reported to be USD 4.2 million. ^28^

The widespread implementation of statin therapy in Germany was opposed by fears of the “medicalization” of the population and references to non-drug interventions such as exercise or a “heart-healthy” diet. Their potential has been known and undisputed for many years. However, the sustainable implementation of lifestyle modifications, for whatever reason, has had little success to date. Germany is one of the countries with the highest LDL cholesterol concentrations in the world; ^29^ the prevalence rates of overweight, obesity and diabetes mellitus are increasing, especially compared to the turn of the millennium. ^30, 31^

Finally, it should not be ignored that lifestyle changes can also be costly. A cost of USD 1,915 per QALY was reported for a guideline-based, structured smoking cessation program ^32^ . The cost of preventing diabetes mellitus through intensive lifestyle modification is likely to be between USD 6,000 and USD 13,000 per QALY ^33–35^ but have also been reported in the order of USD 50,000 per QALY ^36^ although it is ultimately questionable whether the resources required for the implementation and follow-up of such measures would even be available in the German healthcare system. According to strict health economic criteria, lifestyle interventions are therefore not necessarily preferable to the low-threshold prescription of statins.

Clearly, the proportion of people who qualify for statin prescribing depends on which calculator is used to determine risk. We have observed considerable differences between the commonly used risk algorithms, which are partly due to differences in the predicted endpoints. For example, the one used here and validated for Germany ^12^ PCE ^37^ provides the probability of the first occurrence of non-fatal myocardial infarction, fatal coronary heart disease or any stroke, the ESC-SCORE used from 2003 onwards (hereinafter referred to as SCORE 1 ^38^ ), the probability of cardiovascular death. Therefore, depending on the study, the SCORE 1 was usually less than half as high as the risk according to the PCE. ^12, 13^ Progress could be achieved through international harmonization of the most important risk calculators. ^13^ In the meantime, SCORE 1 (cardiovascular death) has been followed by SCORE2 ^39^ whose endpoint has been harmonized with the combined endpoint of the PCE “multiplicative” (page 3250 in reference ^20^ ) and which, in the final analysis, yields hardly any higher risks than the SCORE 1. ^40^ This is because mortality from cardiovascular events is currently significantly lower than in the 1990s. ^40^

The proportion of people qualifying for statin treatment does not exceed 25 percent of the total population in any of our scenarios. This corresponds to the proportion of 22 to 25 percent that was found after recalibration of the risk calculators by Pennells et al. ^13^ is reported. They are even lower than the figure reported by Mortensen ^40^ based on the PCE with a risk threshold of 7.5 percent of the Danish population and in the order of 26 percent according to the recommendations of UK-NICE. ^40^

However, the application of the age-dependent risk thresholds of the SCORE2, as recommended by the ESC, means that only 4 percent of the population and only 1 percent of women qualify for treatment. The reasons for this are the underestimation of risk by the SCORE2 ^40^ and insufficient thresholds for initiating treatment. According to Mortensen et al. ^40^ lowering the risk threshold of the SCORE2 to 5 percent would be roughly equivalent to a risk threshold of 7.5 percent according to PCE. These considerations show that an untested application of the current ESC guideline and its risk thresholds to Germany is not justified.

Measured in terms of healthcare costs, life expectancy in Germany is disappointingly low by international standards, which is attributed to deficits in cardiovascular prevention. ^41^ Comprehensive, low-threshold and early treatment with statins at least has the potential to increase the average life expectancy of the population. This applies above all to the population treated with statins. However, the average life expectancy of the population as a whole is increased to a lesser extent. The associated annual costs range between 0.49 and 1.93 billion euros (Table 2, column 8) and thus between 0.5 and 4 percent of the current pharmaceutical expenditure of around 50 billion euros. At present, it is not possible to estimate whether the implementation of low-threshold primary prevention with statins would actually result in such additional costs. This is because in 2022, EUR 0.841 billion was already spent on lipid-lowering drugs, around half of which was spent on statins, although this also includes an unknown proportion of prescriptions for primary prevention. ^42^

The authors can only mention here that, given limited resources, other, *quoad vitam* obviously less effective but previously reimbursable services may have to be restricted. After all, in the medium term, the prioritization of healthcare services based on key health economic data will also depend on the results of a transparent social discourse that is free from vested interests.

Overall, our data show that the low-threshold prescription of moderately to highly effective statins can be a cost-effective option for the primary prevention of cardiovascular disease. Although age-dependent risk thresholds are plausible, for pragmatic reasons we prefer a risk threshold of 7.5 percent in 10 years, which is in line with US recommendations.

Finally, we emphasize that our work is subject to the limitations of any health economic analysis. It is a simulation based on assumptions that does not take into account uncertain factors such as medical progress (e.g. gene therapy), disease progression, cost trends or patient preferences. The results are therefore merely a schematic decision-making aid for the medical profession and other stakeholders in the healthcare system. The benefits and risks of statin therapy must be weighed up by the practitioner in each individual case, even if the long-term side effect potential of statins is low compared to the clinical benefit. ^7^ Finally, it does not seem opportune to us to merely enshrine the right to statin treatment in law and to ignore other important pillars of primary prevention such as weight reduction, treatment of diabetes mellitus and hypertension. Statin therapy is undoubtedly effective and cost-effective even with a lower global risk than 20 percent in 10 years. The open question is whether society wants to afford such a therapy. What is certain, however, is that “Western” healthcare systems can currently afford much more expensive therapeutic measures without serious concerns about “cost-effectiveness”.

## Data Availability

All data produced in the present study are available upon reasonable request to the authors

## Authors and contributions

AD and WM conceived the study, interpreted the results and critically reviewed the manuscript. AD calculates results. FF prepared database and assumptions für underlying Markov Models and critically reviewed the manuscript. BK and GK critically reviewed that manuscript and made important intellectual contributions. All authors take full responsibility of the whole manuscript.

